# A Comparison of Separate Access versus In-Line Configuration for Continuous Renal Replacement Therapy in VV ECMO

**DOI:** 10.1101/2022.11.16.22282382

**Authors:** Michael Kouch, Adam Green, Solomon Dawson, Christopher Noel, Meghan Gorski, Robert Rios, Nitin Puri

## Abstract

**Objective:** Continuous renal replacement therapy (CRRT) in patients receiving venovenous extracorporeal membrane oxygenation (VV ECMO) can be accessed via separate venous access (SVA) or directly “in-line” within the ECMO circuit. The objective of this study is to compare the efficacy and safety of SVA versus in-line configuration in patients requiring CRRT and VV ECMO.

**Design:** A retrospective review of 16 subjects receiving CRRT while on VV ECMO support.

**Setting:** Adult ICU at a tertiary academic medical institution.

**Patients:** Critically ill adult subjects with severe respiratory failure undergoing percutaneous VV ECMO and CRRT.

**Interventions:** CRRT with venous access via separate temporary hemodialysis catheter versus direct access in-line with the VV ECMO circuit.

**Measurements and Results:** Subject demographics, ECMO cannulation configuration, duration of CRRT, oxygenator and CRRT filter lifespan, number of interruptions, and dialysis blood flow rate were recorded. Five patients received CRRT via SVA and 11 patients via in-line configuration. There was a higher mean number of days on CRRT in the in-line group (7 vs 12 days, p = 0.32). There was no significant difference in oxygenator lifespan (days) (17.1 vs 14.72, p = 0.597), CRRT filter lifespan (days) (1.68 vs 2.15 p = 0.136), or interruptions per 100 CRRT days (10 vs 11.73, p = 0.834) in SVA vs in-line groups. Percentage days with therapeutic anticoagulation (65 vs 68, p =0.859) and initial dialysis blood flow rate (300 vs 310.91 mL/min, p = 0.258) were similar between configurations. SVA was associated with frequent access site manipulation (60% of patients) and catheter site bleeding (40% of patients).

**Conclusions:** CRRT can be delivered via SVA or in-line configuration in patients requiring VV ECMO with similar efficacy. Separate venous access for CRRT may be associated with higher rates of access associated bleeding and need for access manipulation when compared to in-line configuration.

**Key Points:** *Question:* Do separate venous access and in-line configuration for continuous renal replacement therapy (CRRT) in patients requiring VV ECMO have different safety and feasibility profiles? Findings: This retrospective review showed no significant difference in oxygenator lifespan (days) (17.1 vs 14.72, p = 0.597), CRRT filter lifespan (days) (1.68 vs 2.15 p = 0.136), or interruptions per 100 CRRT days (10 vs 11.73, p = 0.834) between separate venous access and in-line configuration groups. While there was no significant difference in mortality (40% vs 72.73%, p = 0.299), separate venous access was associated with frequent access site manipulation (60% of patients) and catheter site bleeding (40% of patients).

*Meaning:* CRRT in patient requiring VV ECMO can be achieved via separate venous access or in-line configuration with similar safety and feasibility specifically regarding oxygenator and filter function.

*Summary Statement:* Continuous renal replacement therapy can be delivered via separate venous access or in-line configuration in patients requiring VV ECMO. Both dialysis access configurations display similar efficacy as described by oxygenator and CRRT filter lifespan, number of CRRT interruptions, and dialysis blood flow rates. Separate venous access for CRRT may be associated with higher rates of access associated bleeding and need for access manipulation when compared
to in-line configuration.

## Introduction

Venovenous extracorporeal membrane oxygenation (VV ECMO) is a potentially life-saving therapy used in patients with reversible causes of respiratory failure refractory to conventional mechanical ventilation. VV ECMO usage continues to increase with improvement in mortality among all indications for extracorporeal life support (ECLS)^1^. Despite improving mortality, complications including acute kidney injury (AKI), infection, thrombosis, circuit mechanical failure, and bleeding are the most frequently encountered adverse events^1-3^. The incidence of AKI ranges from 9.3% to 46% in patients receiving VV ECMO due to different definitions, patient characteristics, and reporting criteria^1,3–4^. Two separate studies showed a pooled incidence of greater than 40% of AKI requiring renal replacement in patients on VV ECMO^3,4^. This was associated with higher mortality and lower rates of mechanical ventilation weaning^4^. Fluid overload is associated with higher mortality and longer duration of ECMO support. Treatment or prevention of fluid overload are the most common indications for initiation of RRT even in the absence of AKI in this patient population^5^.

Renal replacement therapy can be provided via continuous RRT (CRRT), prolonged intermittent renal replacement therapy (PIRRT), intermittent hemodialysis (IHD), and peritoneal dialysis (PD). CRRT is the predominate configuration used in patients on ECMO due to less fluid shifts, better hemodynamic stability, and more precise control of fluid balance^6-8^. Several configurations exist to deliver RRT to the ECMO patient. These include using an in-line hemofilter, an integrated system connecting an RRT device into the ECMO circuit, and a parallel system using separate venous access and RRT machine^9,10^. Choice of RRT configuration varies among institutions and is often dictated by patient characteristics. In a survey of 65 ECMO centers, 50.8% of centers exclusively used a CRRT machine connected to the circuit and 21.5% used in-line hemofilter^5^. While no configuration has been proven to be superior, integrating RRT directly into the ECMO circuit (integrated system) has theoretical risks including air embolism, infection, circuit clotting, shunt within the ECMO system, and high pressures within the RRT system that prevent the machine functioning. This is versus the inherent complications or limitations of obtaining separate vascular access within the parallel system. This study compares the safety and feasibility of an integrated versus parallel system of RRT in patients on VV ECMO for respiratory failure with a focus on complications as well as oxygenator and dialysis filter function and lifespan.

## Materials and Methods

### Statistical Methods

The study consisted of a retrospective observational review of all patients receiving VV ECMO support via peripheral cannulation and simultaneous CRRT at a tertiary academic medical center from April 2020 to November 2021. The retrospective review was approved by Cooper University Hospital Institutional Review Board (IRB# 21-145) on 7/29/2021 under the title “Evaluation of CKRT Configurations During VV-ECMO.” Procedures were followed in accordance with ethical standards of the responsible committee on human experimentation and with the Helsinki Declaration of 1975. Informed consent was waived. Data were retrieved from the electronic medical record (EPIC Systems, Verona, WI USA). Data included demographics, clinical characteristics, venous access complications, need to change or manipulate vascular access, central line associated bloodstream infection (CLABSI), percent of ECMO days with therapeutic anticoagulation, lifespan of the CRRT filter, number of dialysis interruptions per 100 CRRT days, and oxygenator lifespan.

Venous access complications were defined as pneumothorax or arterial puncture resulting from catheter insertion, air embolism, or bleeding at the sight of access requiring ≥ 1 unit packed red blood cell (PRBC). The need to manipulate vascular access included switching of the CRRT access ports, dwelling of tissue plasminogen activator (tPA), or rotation or repositioning of the catheter. This information was obtained from CRRT nursing flowsheets, physician notes, and nursing shift event notes. CLABSI was defined as unexplained bacteremia deemed not to be related to a contaminant without an alternative source of infection. This required ≥ 2 positive blood cultures from 2 different venipuncture sites. Therapeutic anticoagulation was defined as anti-Xa >0.3 or partial thromboplastin time (PTT) > 60 seconds. Data regarding lifespan of CRRT filter and oxygenator were retrieved from physician, nursing, and perfusionist daily notes and flowsheets.

### Inclusion

Patients met inclusion criteria if they were ≥ 18 years old and fulfilled criteria for initiation of VV ECMO and simultaneous CRRT. Inclusion criteria for VV ECMO is described in supplemental Figure 1. All causes of respiratory failure were deemed reversible by the treating physician. Patients were deemed candidates for CRRT based on expert opinion of consulting board certified nephrologists. VV ECMO was performed using Rotaflow centrifugal pumps (Maquet, Rastatt, Germany) with Quadrox membrane gas exchangers (Maquet, Rastatt, Germany). CRRT was performed using Nx stage system I (NxStage Medical, Massachusetts, USA). All cannulations for VV ECMO were performed by medical intensive care physicians^11^. All CRRT venous access procedures were performed by either medical intensive care physicians or critical care fellows under attending supervision. Patients were included into two groups based on CRRT access configuration. Group 1 included separate venous access via temporary hemodialysis (HD) catheter placed in an internal jugular, femoral, or subclavian vein. Group 2 included patients with CRRT access tubing integrated directly in-line with the ECMO circuit as described below. This group will be described as “in-line” with the ECMO circuit but does not include an integrated hemofilter within the ECMO system. Patients who received CRRT for periods of time in each of the two configurations were excluded.

**Figure 1:**
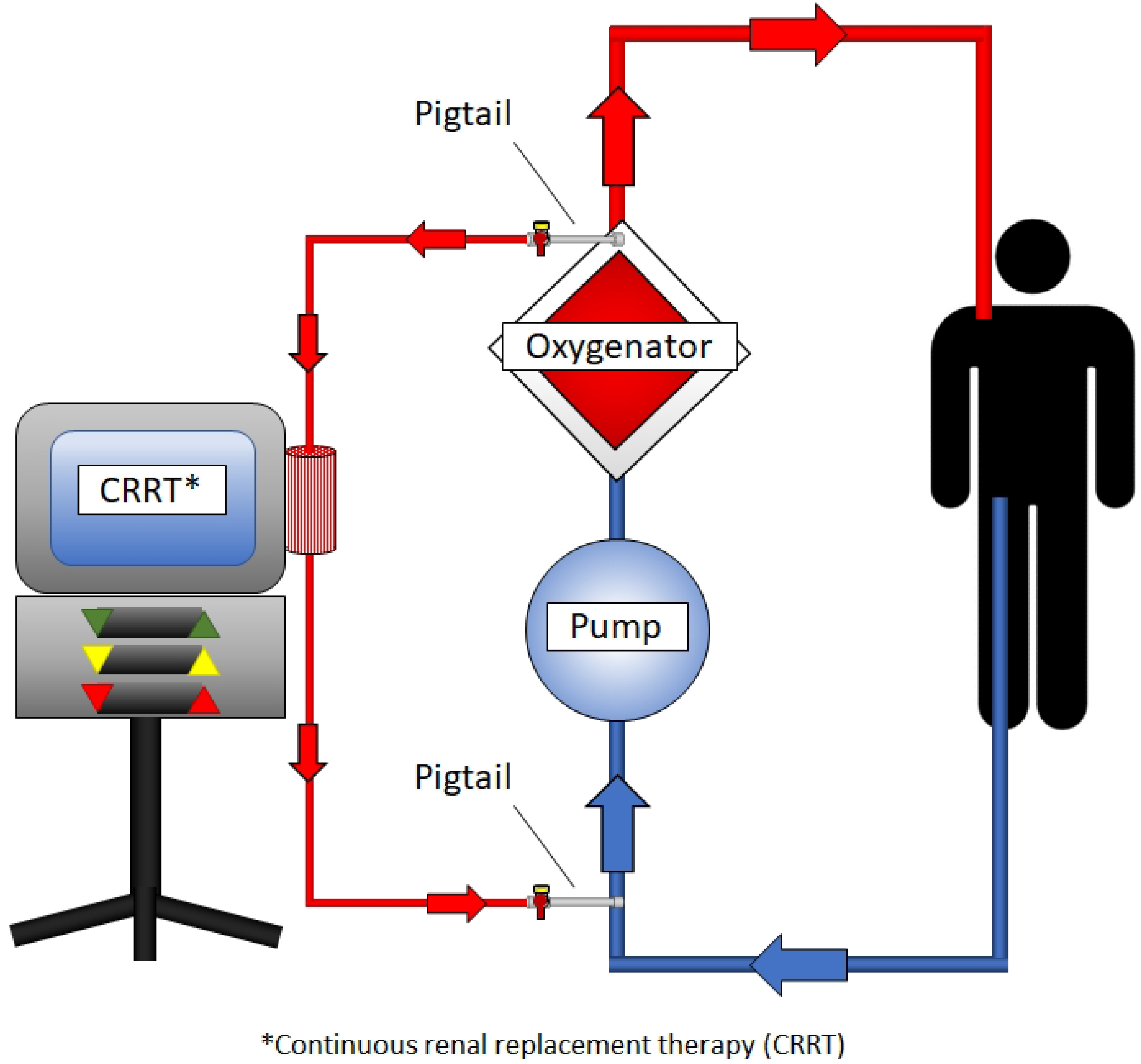
CRRT in-line configuration set up

### CRRT In-Line Configuration

Connection of the CRRT lines to the ECMO circuit was performed as described below and depicted in Figures 1-3. The inflow line delivers blood from the ECMO circuit to the CRRT machine and the outflow line returns blood back to the patient via the ECMO circuit. A custom-made pigtail catheter (Terumo Medical, New Jersey USA) is used for connections as described below. This pigtail catheter consists of 90 durometer pressure tubing with internal diameter of 0.6 inches in an outside diameter of 0.13 inches (Figure 4). The pigtail catheter connects to the oxygenator through the de-airing port on the post-oxygenator side (Figure 2). This pigtail then connects to the CRRT tubing by a three-way stopcock. Because this connection is after the centrifugal pump of the ECMO circuit, it is under positive pressure. The return tubing after the CRRT machine connects to the pigtail catheter which is connected to the pre-pump and pre-oxygenator side of the ECMO circuit (Figure 3). In this configuration, the alarms on the CRRT machine do not need to be adjusted. Because this connection occurs in the negative pressure portion of the ECMO circuit before the centrifugal pump, there is no resistance to trigger high-pressure alarms. The pigtail catheter serves as a resistor against negative pressure alarms from the CRRT machine. All ECMO circuits at this institution have these pigtails bonded to a Leur-lock at these connection sites to avoid the need to clamp the circuit for insertion of the pigtail catheter should this in-line configuration be needed.

**Figure 2:**
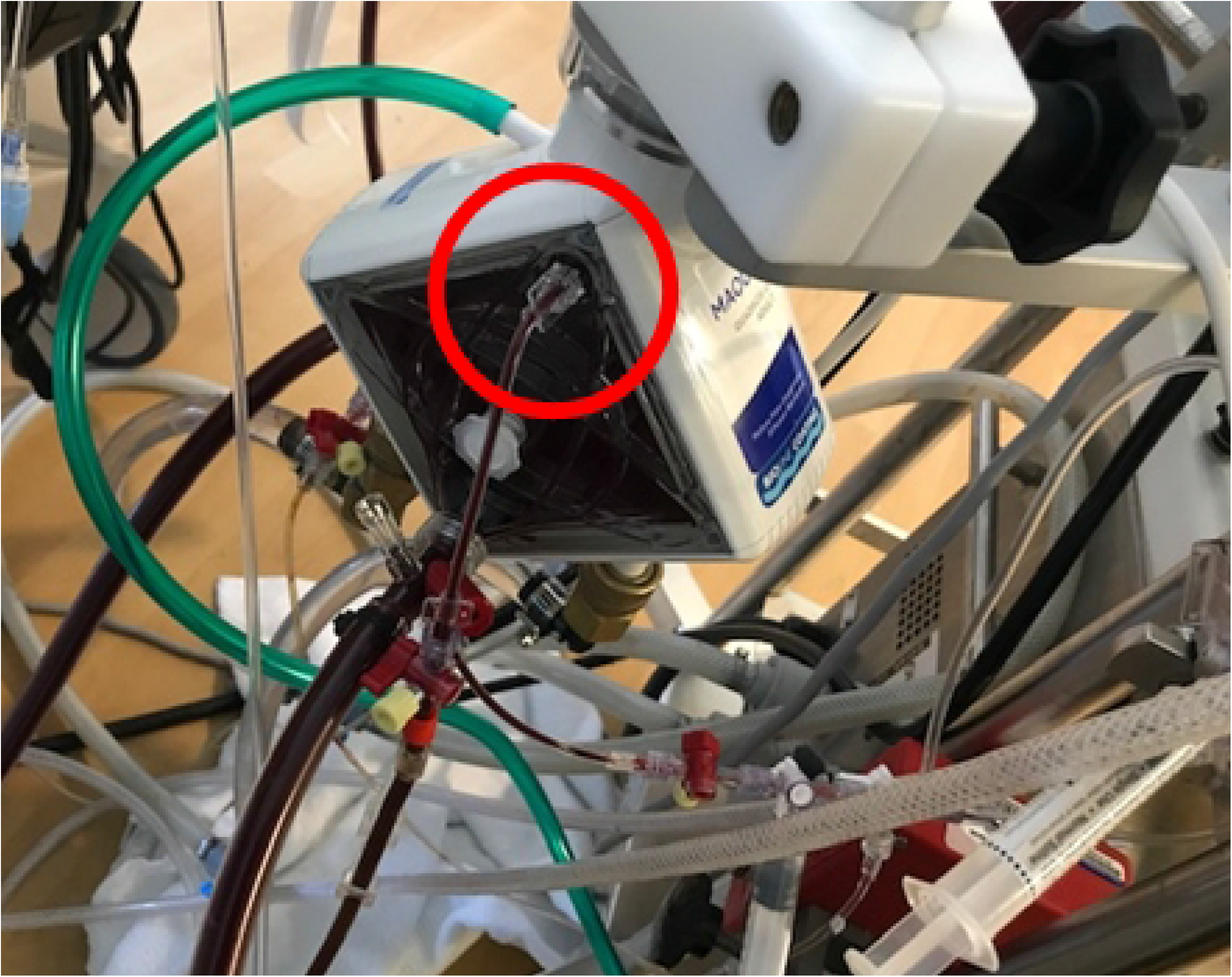
Post-pump and post-oxygenator inflow CRRT-ECMO configuration connection

**Figure 3:**
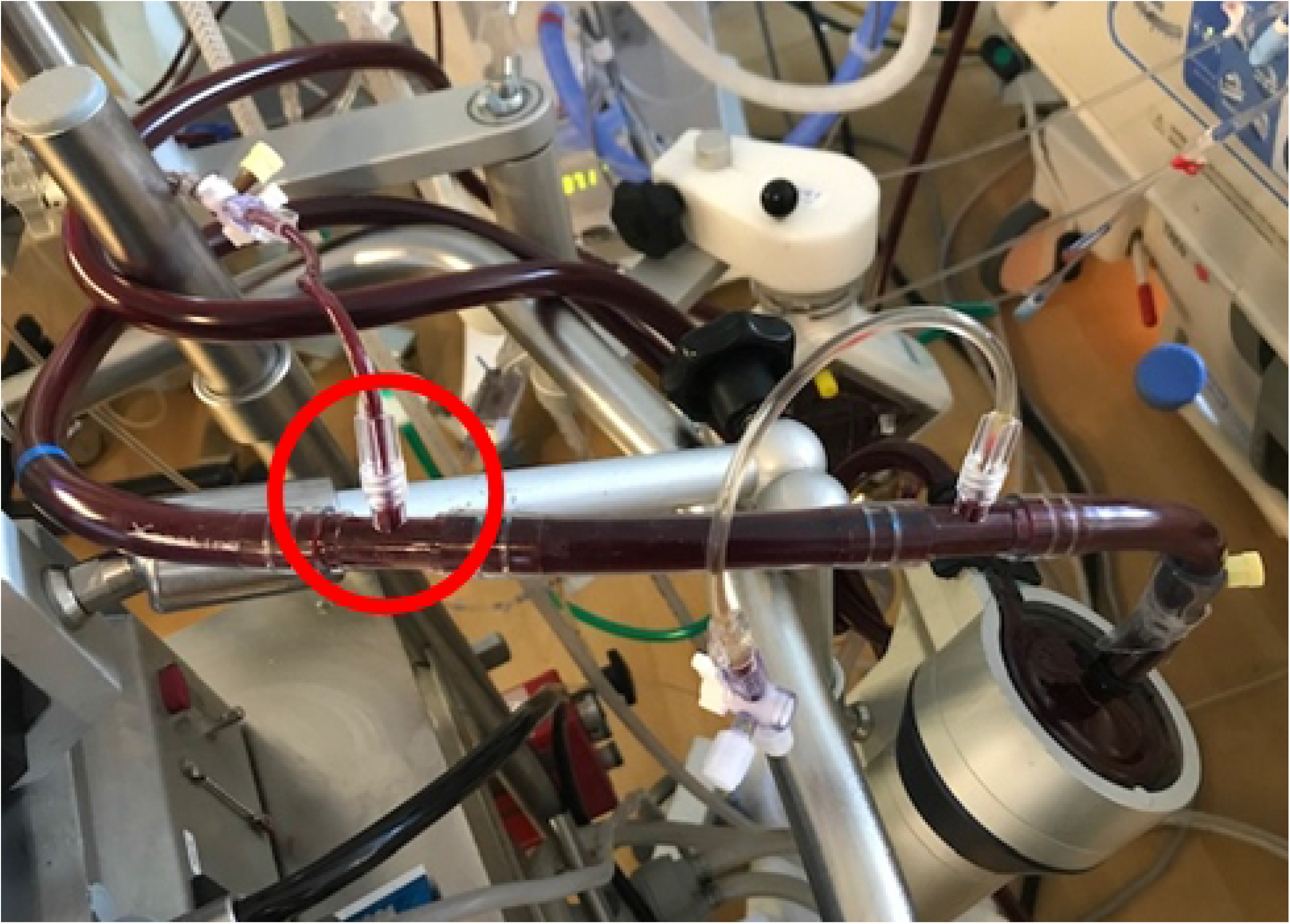
Pre-pump, pre-oxygenator outflow CRRT-ECMO configuration connection

**Figure 4:**
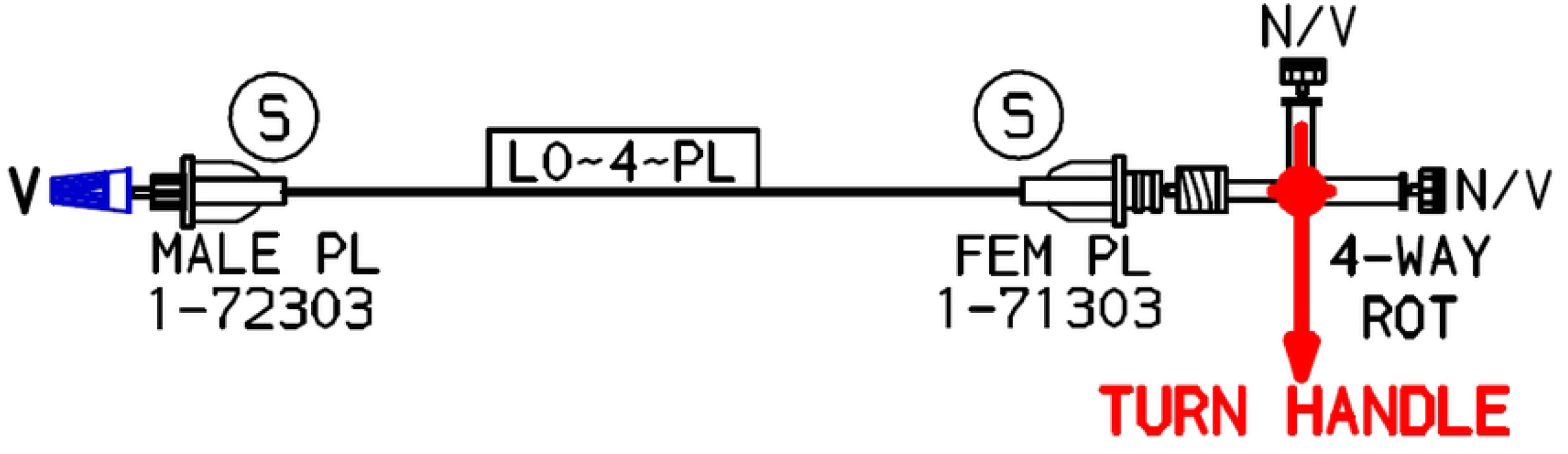
Specialty pigtail connector design

## Results

A total of 20 patients received VV ECMO and concomitant CRRT between April 2020 to November 2021. Five patients were treated with CRRT using separate venous access (SVA) with a temporary hemodialysis catheter, and 11 patients received CRRT via in-line (integrated) configuration with the ECMO circuit. Three patients received a combination of access configurations and were excluded from data analysis. Two of these patients were transitioned from separate access to in-line configuration due to CRRT catheter and machine clotting. The third patient was transitioned from in-line to separate access due to ECMO flow disturbances. Ultimately, this patient was transitioned back to in-line configuration as separate access did not impact the ECMO flow disturbances. Data will be presented as separate access vs in-line configuration. Patient characteristics are shown in Table 1. There was no significant difference in age, BMI, or race between groups. The majority of patients in all groups were diagnosed with respiratory failure due to COVID-19 Pneumonia (80 vs 90.1, p = 1). There was no significant difference in initial CRRT blood flow rates (300 vs 310.91, 0.258) or percentage of ECMO days with successful therapeutic anticoagulation (65 vs 68, 0.859). Mortality rates were also not significantly different (40% vs 72.73%, p = 0.299).

**Table 1:**
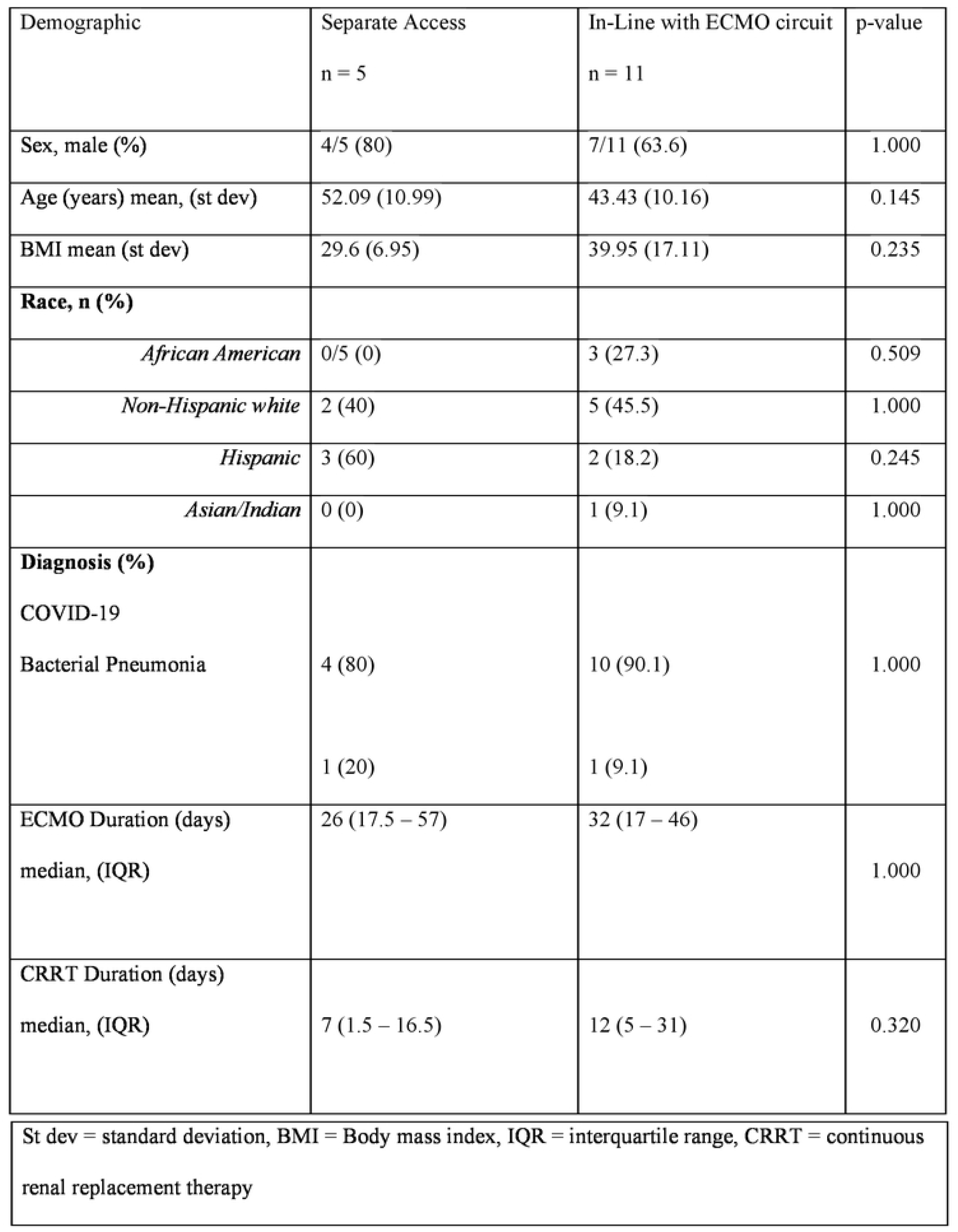
Demographics.

There was no significant difference between groups in oxygenator lifespan in days (17.1 vs 14.72, p = 0.597) or CRRT filter lifespan (1.68 vs 2.15, p = 0.136). There was no significant difference between number of CRRT interruptions or alarms per 100 CRRT days (10 vs 11.73, p = 0.834).

There was no evidence of complications related to venous access including air embolism, unexplained bacteremia, and insertion complications in both groups; data is included in Table 2. Venous access site bleeding and need for catheter manipulation was present in 40% in the SVA group.

**Table 2:** Outcomes.

| Outcome Measure | Separate Access<br>n = 5 | In-Line with ECMO circuit<br>n = 11 | p-value |
| --- | --- | --- | --- |
| CRRT Access Insertion<br>Complications<br>n (%) | 0 | n/a |  |
| CRRT catheter site bleeding<br>requiring transfusion<br>n (%) | 2 (40) | n/a |  |
| Air Embolism<br>n (%) | 0 | 0 |  |
| Unexplained bacteremia<br>n (%) | 0 | 0 |  |
| Catheter Manipulation<br>n (%) | 3 (60) | n/a |  |
| Oxygenator Lifespan<br>(days) (Mean) (st dev) | 17.1 (12.64) | 14.72 (5.41) | 0.597 |
| CRRT Filter Lifespan<br>(days) (Mean) (st dev) | 1.68 (0.76) | 2.15 (0.44) | 0.136 |
| Number of CRRT interrupts or | 10 (22.26) | 11.73 (10.81) |  |
| alarms per 100 CRRT days<br>Median (SD) |  |  | 0.834 |
| % Total ECMO days Xa > 0.3, PTT<br>> 60 (not just days on CRRT) Mean,<br>(st dev) | 65 (37) | 68 (33) | 0.859 |
| BFR (mL/min) of dialysis machine at<br>initiation<br>(st dev) | 300 (0) | 310.91 (30.15) | 0.258 |
| Mortality (%) | 2 (40) | 8 (72.73) | 0.299 |
| CRRT = continuous renal replacement therapy, st dev =. Standard deviation, ECMO = extracorporeal<br>membrane oxygenation, BFR = blood flow rate |  |  |  |

\*Continuous renal replacement therapy (CRRT)

## Discussion

Previous studies have described different configurations for renal replacement therapy in the ECMO population^12-17^. While there is no definitive optimal configuration, previous authors have highlighted the potential risks and benefits of each set up ^12-18^. This retrospective study demonstrates the safety and efficacy of our specific in-line configuration in the VV ECMO population when compared to using separate venous access for CRRT. Both separate venous access and in-line configuration groups showed similar oxygenator and CRRT filter lifespan without evidence of severe adverse events such as air embolism or venous access insertion complications.

Prior literature shows similar efficacy and safety of in-line or integrated configurations when focusing on pressure alarms and hemolysis^13,15,16^. An in-line CRRT connection was first described 7 patients on VA ECMO with dialysis blood flow rates reaching 150-200 mL/min^15^. Likewise, in-line configuration in patients receiving ECLS with CRRT inflow originating from the arterial (ECMO return) end of the ECMO circuit and the CRRT outflow to the pre-pump side of the ECMO circuit has also been described with shorter delay to initiate dialysis and no thromboembolic or septic complications^16^. While our study shows no significant difference in CRRT filter life span between groups, previous literature describes longer CRRT filter lifespan when CRRT is introduced into the ECMO circuit (in-line or integrated configuration) in a pediatric patient population^17^. While comparison of these two populations is limited, we hypothesize that the filter lifespan was influenced by the lower blood flow rate in the pediatric population compared to our study.

Literature regarding mean oxygenator lifespan is rather limited. Nonetheless, oxygenator lifespan was similar in both groups in our study and twice as long as previously described^18^. This is particularly significant due to the high percentage of patients with COVID 19 infection and the increased risk of thrombosis and coagulopathy in this population when receiving ECMO support^19^. CRRT filter lifespan was longer than expected in each group as prior literature has shown decreased filter lifespan in critically ill patients with COVID 19^20^. We hypothesize the longer lifespan of each device to be related to the high initial CRRT blood flow rates (BFR) (300 vs 310.91, p = 0.258) even with only moderate percentage of days of achieved therapeutic anticoagulation in each group (65 vs 68, p = 0.859)^21,22^.

Furthermore, in our study the in-line group was associated with less CRRT alarms or interruptions per 100 CRRT days (10 vs 11.73, p = 0.834). While this did not meet statistical significance, the clinical importance of time on CRRT for volume removal is likely relevant. This outcome is similar to prior literature describing successful management of CRRT pressure alarms using direct connection of CRRT to the ECMO circuit without manipulation of CRRT alarm settings or modifying ECMO settings by using three different configuration options in which the CRRT inflow catheter was connected either between the ECMO pump and oxygenator or before the ECMO pump^13^. Prior authors have highlighted the risk of air embolism if access was connected post oxygenator. Our configuration mitigates this risk as the 3-way stopcock and Leur lock is connected the post oxygenator side of the circuit in all patients during the initiation of ECMO. Therefore, clamping and connection of the Leur lock is not needed if the patient would require CRRT after initiation of ECMO.

We hypothesize the relative efficacy and safety of this configuration is driven by the connection of the Leur lock to the ECMO circuit in all patients cannulated for VV ECMO. The CRRT line returns to the negative pressure portion of the ECMO circuit with pressures ranging from -20 to -100 mmHg. Prior authors have commented on the risk of low negative pressure alarms stopping the CRRT machine, hemolysis, or embolism^12,13^. The Leur lock serves as a resistor to the negative pressure, thereby preventing alarms; blood flow return to the pre-oxygenator side of the circuit lessens the risk of systemic embolic events.

The in-line configuration group had a higher mortality rate than those in the separate venous access and combination groups (40 vs 72.72, p = 0.299). While this did not meet statical significance nor is this study powered to show differences in mortality, it is important to highlight the higher percentage of COVID 19 patients in the in-line configuration group. Early pandemic data showed high mortality in COVID 19 patients supported with ECMO^23^. While later cohort studies have shown no difference in outcomes between patients with COVID 19 versus alternative causes of ARDS supported with ECMO, there is an increased survival for patients in the late stage of the pandemic compared of the early stage (before May 2020)^23,24^. Because this study includes both stages of pandemic, we advise against the assumption that increased mortality in the in-line configuration group is a result of CRRT configuration due to the many confounding variables. It is also possible that sicker patients were included in the in-line configuration group due to the potential limitations in vascular access in the sicker cohort.

Limitations of this study include those inherent to the retrospective nature and the small sample size. Duration of filter or oxygenator lifespan is impacted by death and decannulation. If a filter or oxygenator is changed shortly before either event, the true duration of each device would be underestimated. Markers of hemolysis are absent in our study. Likewise, dialysis efficiency data such as dialysis recirculation fraction is not recorded. Transfusion requirements are often multifactorial and cannot be attributed vascular access site bleeding alone. While separate vascular access site manipulation is recorded in nursing flowsheets, data for manipulation of connection points for the in-line setup are absent for comparison. This study focuses on one configuration using a specific pigtail and Leur lock connection. It cannot necessarily be extrapolated to alternative in-line configurations. Patients on venoarterial ECMO were not included in this study, and this data cannot be extrapolated to that population. Future prospective studies may include dialysis efficacy, hemolysis parameters, and time to initiate dialysis.

## Conclusion

CRRT can be delivered via separate venous access or in-line configuration in patients requiring VV ECMO. Both dialysis access configurations display similar efficacy and safety. Separate venous access may be associated with higher rates of access associated bleeding and need for access manipulation.

## Data Availability

Data cannot be shared publicly because of private information. Data are available from the Institutional Data Access / Ethics Committee (contact via email) for researchers who meet the criteria for access to confidential data.

BFR: blood flow rate
BMI: Body mass index
CRRT: continuous renal replacement therapy
ECMO: extracorporeal membrane oxygenation
IQR: interquartile range
St dev: standard deviation

## Notes

<u>Financial Support:</u> Support was provided solely from institutional and/or departmental sources.

<u>Conflicts of Interest:</u> The authors declare no conflicts of interest in connection with the study reported

### Competing Interest Statement

The authors have declared no competing interest.

### Funding Statement

The author(s) received no specific funding for this work.

### Author Declarations

The retrospective review was approved by Cooper University Hospital Institutional Review Board (IRB# 21-145) on 7/29/2021 under the title “Evaluation of CKRT Configurations During VV-ECMO.” Procedures were followed in accordance with ethical standards of the responsible committee on human experimentation and with the Helsinki Declaration of 1975. Informed consent was waived.

